# Proteome Wide Association Studies of LRRK2 variants identify novel causal and druggable for Parkinson’s disease

**DOI:** 10.1101/2023.01.05.23284241

**Authors:** Bridget Phillips, Daniel Western, Lihua Wang, Jigyasha Timsina, Yichen Sun, Priyanka Gorijala, Chengran Yang, Anh Do, Niko-Petteri Nykänen, Ignacio Alvarez, Miquel Aguilar, Pau Pastor, John C. Morris, Suzanne E. Schindler, Anne M. Fagan, Raquel Puerta, Pablo García-González, Itziar de Rojas, Marta Marquié, Mercè Boada, Agustin Ruiz, Joel S. Perlmutter, Dominantly Inherited Alzheimer Network (DIAN) Consortia, Laura Ibanez, Richard J. Perrin, Yun Ju Sung, Carlos Cruchaga

**Affiliations:** Department of Psychiatry, Washington University School of Medicine, St. Louis, MO 63110, USA; NeuroGenomics and Informatics Center, Washington University School of Medicine, St. Louis, MO 63110, USA; Hope Center for Neurological Disorders, Washington University School of Medicine, St. Louis, MO 63110, USA; Division of Biostatistics, Washington University; Memory Disorders Unit, Department of Neurology, University Hospital Mutua Terrassa, Terrassa, Spain; Unit of Neurodegenerative diseases, Department of Neurology, University Hospital Germans Trias i Pujol and The Germans Trias i Pujol Research Institute (IGTP) Badalona, Barcelona, Spain; Department of Neurology, Washington University School of Medicine, St. Louis, MO 63110, USA; Department of Pathology and Immunology, Washington University School of Medicine, St. Louis, MO 63110, USA; Ace Alzheimer Center Barcelona - Universitat Internacional de Catalunya, Barcelona, Spain; Networking Research Center On Neurodegenerative Diseases (CIBERNED), Instituto de Salud Carlos III, Madrid, Spain

**Author notes:** Correspondence to: Carlos Cruchaga, PhD, Full address: 4444 Forest Park Ave, Washington University School of Medicine, St. Louis, MO 63110, USA.

**Keywords:** Parkinson’s disease, LRRK2, Proteomics, PheWAS, GRN

## Abstract

Common and rare variants in the *LRRK2* locus are associated with Parkinson’s disease (PD) risk, but the downstream effects of these variants on protein levels remains unknown. We performed comprehensive proteogenomic analyses using the largest aptamer-based CSF proteomics study to date (7,006 aptamers (6,138 unique proteins) in 3,107 individuals). We identified eleven independent SNPs in the *LRRK2* locus associated with the levels of 26 proteins as well as PD risk. Of these, only eleven proteins have been previously associated with PD risk (e.g., GRN or GPNMB). Proteome-wide association study (PWAS) analyses suggested that the levels of ten of those proteins were genetically correlated with PD risk and seven were validated in the PPMI cohort. Mendelian randomization analyses identified five proteins (GPNMB, GRN, HLA-DQA2, LCT, and CD68) causal for PD and nominate one more (ITGB2). These 26 proteins were enriched for microglia-specific proteins and trafficking pathways (both lysosome and intracellular). This study not only demonstrates that protein phenome-wide association studies (PheWAS) and trans-protein quantitative trail loci (pQTL) analyses are powerful for identifying novel protein interactions in an unbiased manner, but also that *LRRK2* is linked with the regulation of PD-associated proteins that are enriched in microglial cells and specific lysosomal pathways.

## Introduction

Leucine-rich repeat kinase 2 (*LRRK2*) gene mutations can cause Mendelian Parkinson’s disease (PD). At the same time, common variant in this locus are some of the most significant PD risk variants. One of the largest PD risk genome-wide association studies (GWAS) to date^1^ found multiple independent signals in the *LRRK2* gene. Subsequent studies^2^ examined the association between the common variant (SNP) rs76904798 (chr12:40220632:C:T) with *LRRK2* RNA expression and found that the minor allele (T) was associated with increased expression levels of *LRRK2*. Like most of the neurodegenerative diseases, PD is a complex disorder with several, and sometimes independently interconnected, pathways encompassing multiple proteins contributing to disease pathogenesis. Identifying those pathways, as well as the included proteins and protein interactions, is instrumental to decipher the pathobiology of the disease.

A recent study^3^ leveraged the cerebrospinal fluid (CSF) proteome to identify the causal proteins under the PD loci by performing cis-protein quantitative trait loci (pQTL) and Mendelian randomization (MR). A limitation of using cis-signals is the non-assessment of potential protein-protein interaction. We recently demonstrated that common variants in *MS4A4A* are a trans-pQTL for CSF soluble TREM2 (sTREM2) levels and are associated to Alzheimer disease (AD) risk,^4^ indicating that MS4A4A modifies risk for AD by modifying TREM2 biology, and that MS4A4A is a potential therapeutic target. This led to several active clinical trials targeting MS4A4A to treat AD. Along the same lines, we performed unbiased multi-tissue pQTL mapping and identified hundreds of cis and trans pQTLs. These pQTLs were later used to identify causal and druggable targets for AD, PD, frontotemporal dementia (FTD), amyotrophic lateral sclerosis (ALS), and stroke.^5^ Together, all these findings strongly support the use of pQTL analyses for the identification of new causal proteins, causal pathways, and potential therapeutic targets.

Here, we performed a trans-pQTL study to identify proteins associated with *LRRK2* variants using the largest aptamer-based CSF proteomics study to date (7,006 aptamers (6,138 unique proteins) in 3,107 individuals), and additional mediation and pathway analyses to understand the downstream effects of *LRRK2* variants (Fig. 1). We discovered that a majority of the identified proteins were enriched in the endolysosomal pathway and immune function.

**Figure 1.**
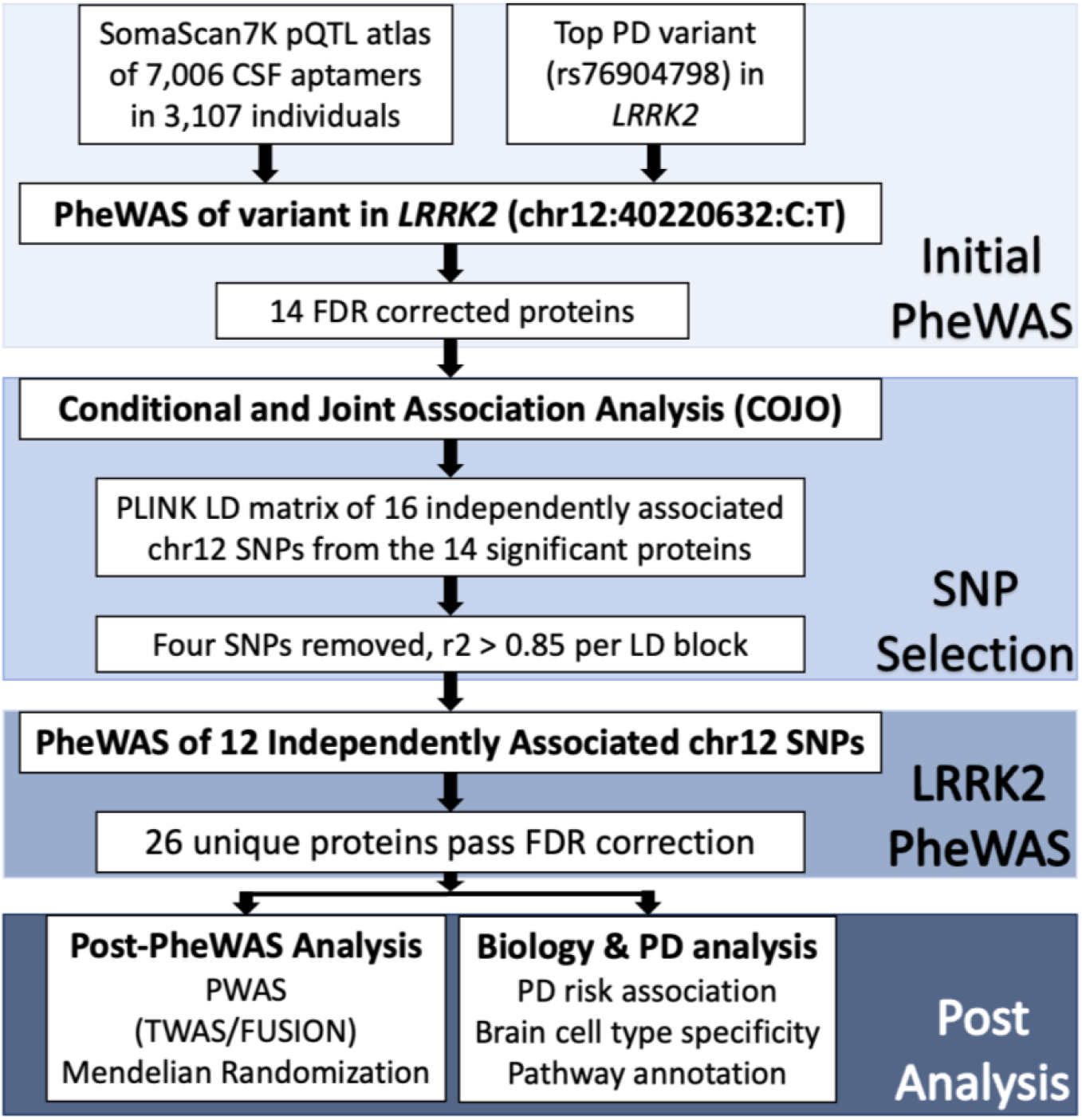
Workflow and study design. The workflow of the LRRK2 loci associated protein analysis with initial PheWAS, SNP selection, *LRRK2* PheWAS, fine mapping of significant aptamers, and analysis of proteins in terms of biological and PD significance.

## Materials and methods

### Study design

We performed phenome-wide association studies (PheWAS) of the sentinel SNP in LRRK2 (rs76904798) using a CSF protein QTL atlas that included more than 3,000 samples to identify causal and druggable proteins for Parkinson disease. We performed conditional analyses (GCTA-COJO) in this region, using those proteins, to identify additional independent SNPs modifying protein levels and PD risk. Next, we leveraged novel statistical approaches to identify genetically associated protein levels (Proteome-wide association studies (PWAS)) and causal proteins (MR). Finally, we performed pathway and cell-type enrichment analyses to determine the biological processes that these proteins are involved in, which major cell-types they are expressed in, and how they interact between each other.

### Cohorts

Protein QTL was performed in a total of 3,107 unrelated, Non-Hispanic White samples recruited from six different cohorts: Dominantly Inherited Alzheimer Network (DIAN), the Charles F. and Joanne Knight Alzheimer Disease Research Center (Knight ADRC), Barcelona-1, Alzheimer’s Disease Neuroimaging Initiative (ADNI), Ace Alzheimer Center Barcelona (Fundació ACE), and Parkinson’s Progression Markers Initiative (PPMI) (Supplementary Table 1). Description of these datasets, and protein QC and processing have been previously described.^6^ DIAN is a longitudinal study of dominantly inherited AD mutation carriers and their family members including asymptomatic and mild to moderate AD. Knight ADRC Memory and Aging Project (MAP) is a longitudinal study of cognitive functioning in persons as they age, with the goal to advance AD research. Barcelona-1 is a longitudinal observational study of dementias such as AD dementia (ADD) and mild cognitive impairment (MCI) at the University Hospital Mutua de Terrassa, Barcelona, Spain. ADNI is a multisite study that aims to develop biomarkers that will improve clinical trials for the prevention and treatment of AD. PPMI is a longitudinal observational study of participants with and without PD, with comprehensive clinical and imaging data and biological samples aimed at identifying markers of disease progression for use in clinical trials of therapies to reduce progression of PD disability.

### Phenome-wide association study and conditional analyses

Proteins were measured using the SomaLogic aptamer-based SOMAscan platform. DIAN, Knight ADRC, Barcelona-1, ADNI, and Fundació ACE CSF samples were measured using SomaScan7K, which measured 7,006 aptamers in 6,139 protein targets. PPMI generated the CSF proteomic data using SomaScan5K and 4,785 aptamers in 4,131 protein targets were measured. Genotyping and protein measurements have been described in detail elsewhere.^5^ There were 3,726 samples, with hg38 GWAS and CSF proteomic data available, included in this study. After genetic principal component analysis (PCA) using the 1000 Genome to select European ancestry, 3,328 samples remained. After removing individuals with cryptic relatedness through identity by descent (IBD) (PIHAT≥0.25), 3,107 samples remained. pQTL linear regression was then performed using PLINK^7^ with the normalized protein levels,^5^ adjusting by age, sex, the first ten principal components, and technical variables (genotyping array). For PD risk, we used the latest published GWAS, which included 2,525,897 total samples in the PD GWAS.^8^

We first determined what proteins were associated with the sentinel SNP on the LRRK2 locus (rs76904798; chr12:40220632:C:T) by performing a PheWAS for rs76904798. Multiple test correction was performed at FDR 5% correction using R package FDRestimation.^9^ We then performed conditional and joint association analysis (GTCA-COJO)^10,11^ on each significant aptamer to identify independently associated SNPs (Supplementary Table 2) on this locus. SNPs with *r*^*2*^>0.85 per LD block were pruned to keep independently associated SNPs without non-random association. Pearson correlation was then performed using the R package corrplot^12^ to determine correlation between proteins.

### Proteome-wide Association Study

A Proteome-wide Association Study (PWAS) was performed to identify associations between Parkinson’s disease risk and protein levels. PWAS^13^ can identify proteins potentially involved in Parkinson’s disease etiology by examining genetically regulated association between proteome expression and PD status. Whereas these approaches were built using only cis signal expression data of the transcriptome, we modified the FUSION^13^ framework to include proteome expression weight files containing both cis and trans signals of the genes. Age, sex, the first ten genetic principal components, and genotyping array were included as covariates in the protein weight calculation model of the PWAS. The functional GWAS association statistic was calculated using the FUSION gusevlab GitHub R script^13^ and protein weights of signals on chromosome 12. FUSION post-processing was then performed to identify conditionally independent associated features by analyzing significant associations within a locus window boundary of 100,000 bp. Three strictly non-overlapping loci were found with the second locus on the 39 to 41.5 Mb region of chromosome 12.

### Validating protein level association with the PPMI cohort

PPMI used DNA samples from blood or saliva for genetic mutation testing of *LRRK2* (p.G2019S and p.R1441G), *GBA* (p.N370S), and *SNCA* mutations. PPMI generated the proteomic data on 1,075 samples using the aptamer-based SomaScan5K platform. A total of 4,783 aptamers remain after QC: outliers were removed by 1.5 IQR threshold, aptamer and individual call rate <65%, and aptamer and individual call rate <85%. There were 917 samples present with PPMI phenotypes including 185 healthy control, 545 PD, and 187 prodromal samples. Only samples with proteomic and clinical data were included in the analyses. Linear regression was performed on the normalized protein levels of PPMI using PLINK^7^ adjusted by age, sex, and cohortArray. Differential protein levels analyses were performed with control samples compared to case, prodromal, and mutation samples.

### Mendelian Randomization

Mendelian Randomization (MR) was performed using the summary statistics of each of the significant aptamer pQTLs generated in here, and the PD GWAS^8^ to investigate the potential causal effect of the proteins on Parkinson’s disease. Using the R package TwoSampleMR,^14,15^ 21 aptamers from 17 proteins had independent genome-wide significant SNPs (*P*<5×10^−8^) present in the pQTL and the PD GWAS. IVW (inverse variance-weighted) p-values, or Wald ratio p-values for one SNP MR, were reported. Since *LRRK2* is in a pleiotropic region and to avoid potential false positives, we performed the analyses again after removing the *LRRK2* locus (39-41.5 Mb region) and then performing MR analyses with cis-pQTLs only.

### Protein-protein interaction and Pathway Analysis

Pathway and interaction analyses were performed using STRING^16^, EnrichGO^17^, DisGeNET^17^, and GeneMANIA.^18^ STRING was performed by entering the 26 genes that code for the proteins identified in this study together with *LRRK2* for gene interactions. EnrichGO was performed with 6,138 SomaScan7K genes as background and 27 genes of interest (26 proteins and *LRRK2*). Gene-disease enrichment was performed using DisGeNET of the 27 genes and 6,138 SomaScan7K genes. Network analysis was performed using GeneMANIA with the default 20 affiliated genes in a network with the 26 genes and *LRRK2*.

### Brain cell enrichment analysis

Cell-type enrichment analysis was performed on the 5,706 genes in CSF SomaScan7K with cell-type data as background and the associated proteins as the target set. Brain cell-types were determined using transcriptome expression data^19^ in microglial/macrophage, endothelial, mature astrocyte, neuronal, and oligodendrocyte cells isolated from human brains. Gene expression data was added for all the samples per gene, and percentiles were calculated per cell-type. If one cell-type had more than 50% of the sum-total expression of a gene, then that gene was considered as cell-type specific. Fold change (FC) was calculated using the ratio between cell percentage for each protein associated to *LRRK2* and the SomaScan7K background set.

## Data availability

Proteomic, pQTL, and raw data from the Knight ADRC participants are available at the NIAGADS and can be accessed at https://www.niagads.org/knight-adrc-collection.

Data from the DIAN cohort is available to qualified investigators and can be requested at https://dian.wustl.edu/our-research/for-investigators/diantu-investigator-resources/dian-tu-biospecimen-request-form/.

ADNI and PPMI proteomic data can be found at: https://adni.loni.usc.edu/ and, https://www.ppmi-info.org/respectively.

## Results

### Proteins associated with *LRRK2* Parkinson’s disease risk variant

To identify proteins associated with *LRRK2* variants, we examined the largest CSF pQTL dataset to identify proteins associated with the sentinel variant in this locus: rs76904798. PheWAS analyses identified 14 proteins in the pQTL atlas with protein levels associated to this variant (C1QTNF1, CD63, CD68, ENTPD1, GPNMB, GREM2, GRN, HLA-DQA2, ITGB2, NIPAL4, OLR1, SDCBP2, TLR3, and TMEM106A). Then, we performed conditional analyses to identify independent signals in this region for these proteins. We identified a total of 15 SNPs associated with these proteins and 12 SNPs were not in LD (r^2^<0.85). Next, we performed a PheWAS of the 12 independent SNPs and identified 12 additional proteins (AGFG2, CA1, CHIT1, DNAJC15, EID3, FCGR1A, FEV, FTL, GAA, LCT, LGALS9, and SRI) (Fig. 2A; Supplementary Fig. 1; Supplementary Table 3). Overall, 27 unique proteins were found to be associated with 12 independent SNPs in the *LRRK2* locus (Fig. 2B; plot made using Gu et al.^20^ Circlize).

**Figure 2.**
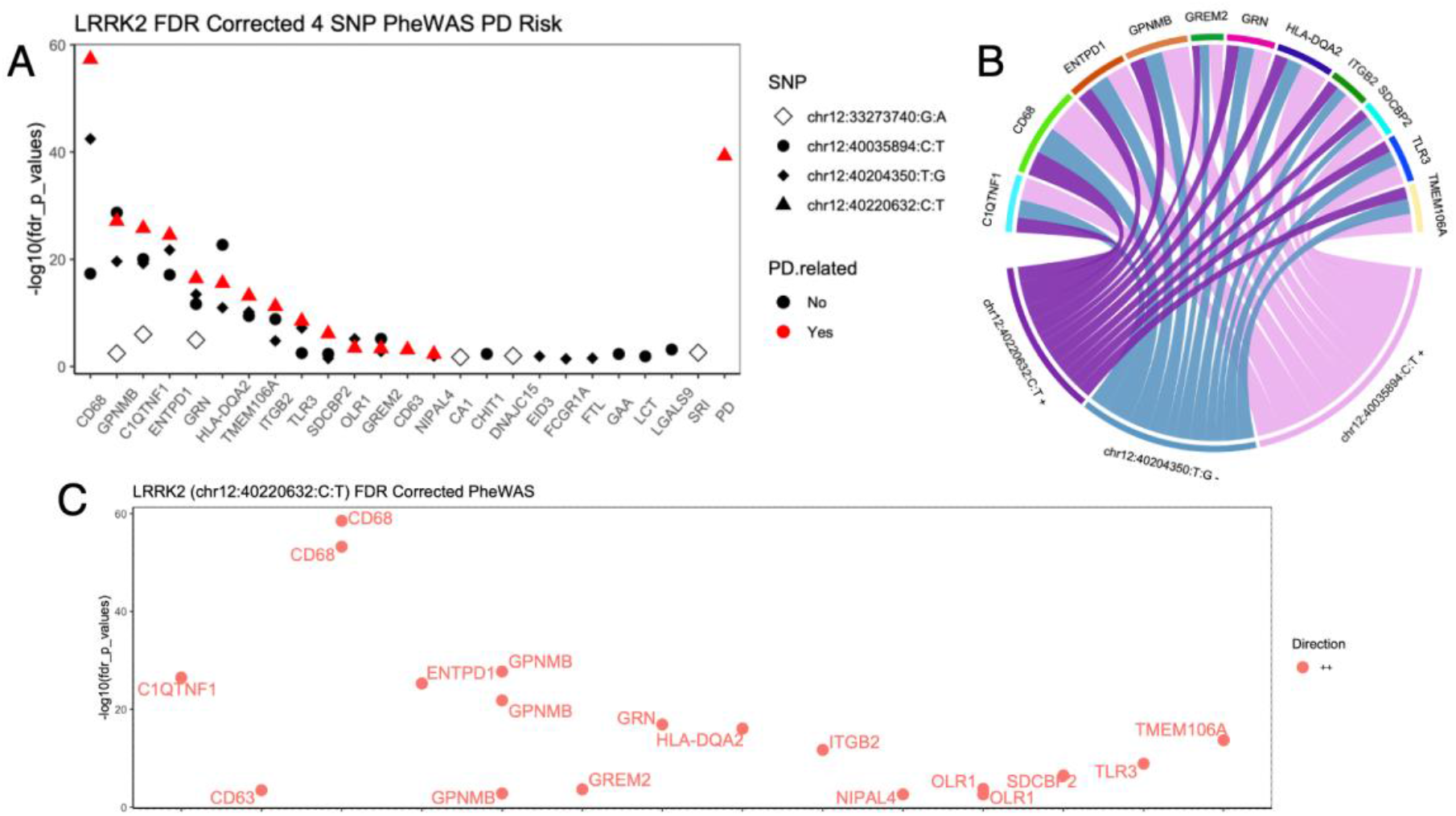
PheWAS and PWAS of LRRK2 variants. (**A**) *LRRK2* PheWAS plot of 24 CSF proteins over four SNPs. Four SNPs shown to display the highest number of proteins with lowest number of SNPs. The PD risk SNP is highlighted red. X-axis is proteins sorted by chr12:40220632:C:T p-values and y-axis is FDR corrected - log10(p-value). For proteins with multiple aptamers, the highest p-value is represented. (**B**) Circos plot of proteins in chr12:40220632:C:T, chr12:40220632:C:T, and chr12:40035894:C:T. Width of connection by SNP effect size. All 14 proteins in chr12:40220632:C:T had positive effect size and the 16 proteins in chr12:40035894:C:T had negative effect size. (**C**) LRRK2 PheWAS plot of 14 CSF proteins (18 aptamers). All FDR passing aptamers had positive chr12:40220632:C:T direction (light red). FDR corrected -log10(p-value) y-axis and x-axis is entrez gene ID.

Multiple proteins (such as GPNMB, CD68, OLR1, and FTL) were measured with more than one aptamer and showed high correlation (*R*>0.9). In addition, GRN and C1QTNF1 (*R*=0.88) showed high correlation. The remaining 20 proteins showed lower correlation (*R*<0.7). (Supplementary Fig. 2).

Eleven (GRN, GPNMB, HLA-DQA2, CD63, CD68, EID3, ENTPD1, GAA, LCT, SRI, and TLR3) of the 26 proteins have been reported to be implicated in Parkinson’s disease pathogenesis association in previous studies (Supplementary Table 4). Six proteins (AGFG2, C1QTNF1, CHIT1, DNAJC15, FTL, and OLR1) have been implicated in AD, ALS, or stroke, but not in Parkinson’s disease. The rest (9) are novel proteins, not implicated with PD or neurodegeneration, such as GREM2, ITGB2, SDCBP2, and TMEM106A.

### Genetically differentially expressed proteins

Then, we integrated the protein QTL with the latest GWAS for PD risk, under the TWAS/FUSION^13^ framework to identify genetically-driven differential expressed proteins in PD (Fig. 2C). This analysis identified eight proteins (HLA-DQA2, GRN, GPNMB, CD68, C1QTNF1, ENTPD1, SDCBP2, and TLR3) with differentially abundant protein levels in PD cases compared to controls, as two (ITBG2 and TMEM106A) with suggestive association (Supplementary Table 5). Higher levels in PD cases compared to controls were predicted for all proteins.

To validate these results, we used the PPMI data to determine if the levels of the PWAS-significant proteins were significantly different between controls, sporadic, PD cases, or individuals with autosomal dominant PD mutations (*LRRK2*+, *GBA*+, *SNCA*+; prodromal cases). Of the 26 proteins (31 aptamers) identified in the initial analyses, 25 had PPMI protein data measure in 741 PPMI samples and diagnosis. We found 16 proteins had significantly different protein levels between controls and cases, controls and prodromal samples, and/or control and mutation carriers (Supplementary Fig. 3; Supplementary Table 6), included the on ones predicted by the FUSION approach: HLA-DQA2, GRN, GPNMB, ENTPD1, ITBG2, and TMEM106A were dysregulated in PD cases, as well as even at the prodromal stages.

### Identifying causal proteins for PD risk

Mendelian randomization was performed to estimate the causal effect of Parkinson’s disease exposure through links between the PD GWAS^8^ and pQTL summary statistics. Our results identified seven proteins, out of the 26, to be causally associated to PD risk (SDCBP2, GPNMB, CD68, LCT, ENTPD1, ITGB2, and C1QTNF1) (Table 1). To address the horizontal pleiotropy of the *LRRK2* locus, we performed MR analyses by removing all the variants in this locus. GRN, GPNMB, HLA-DQA2, CD68, and LCT remained significant. To further confirm that pleiotropy did not cause false positive findings, MR analyses were performed including only cis-pQTLs, with identical results as the previous analyses. These results indicate that GRN, GPNMB, HLA-DQA2, CD68, LCT, and potentially ITGB2 are part of the causal pathway for PD (Table 1).

**Table 1.**
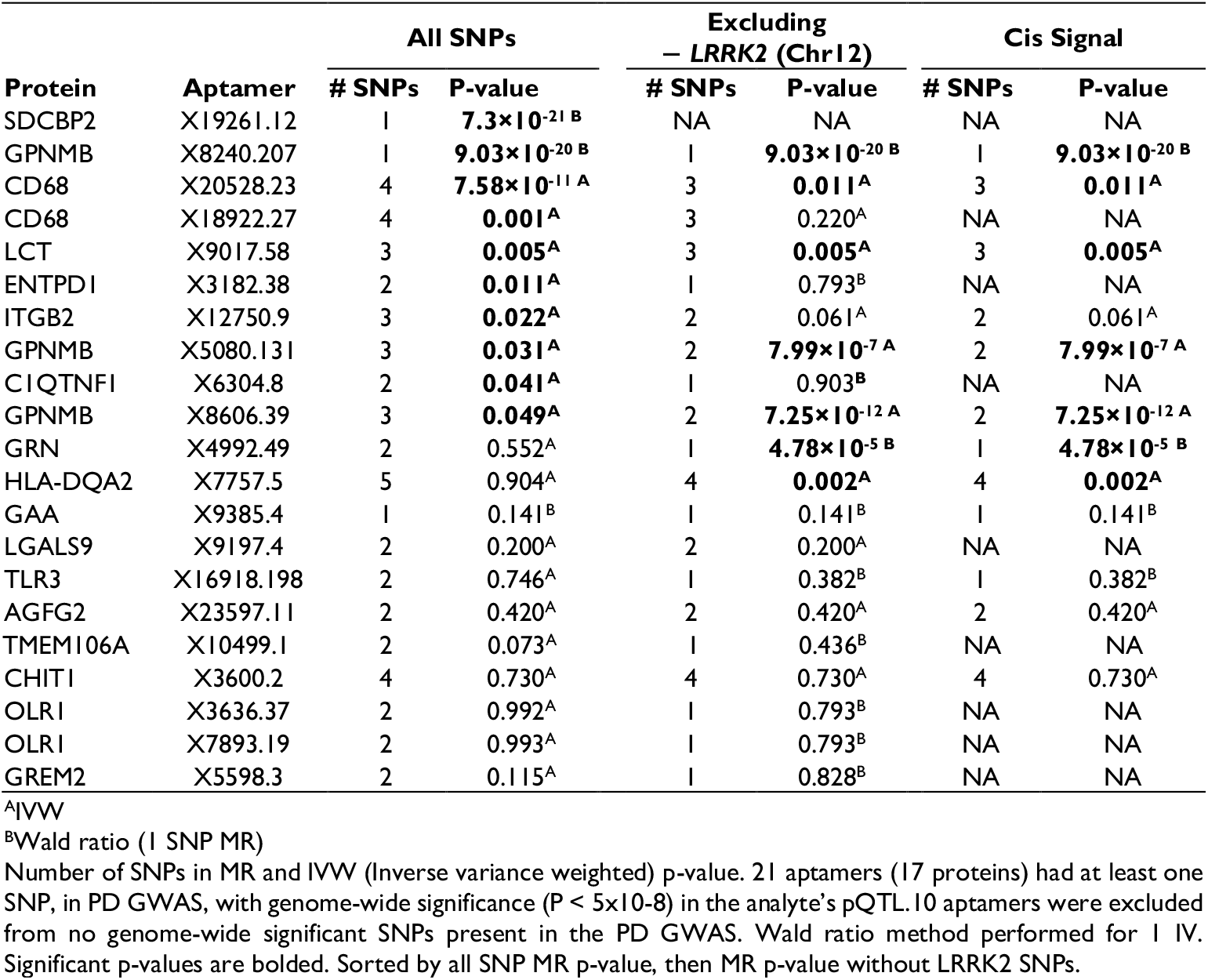
Mendelian Randomization (MR) results of LRRK2 associated proteins for being causal for PD

### Biological insights of the proteogenomic analyses

#### Interaction and Pathway Analysis

Pathway analysis was performed to understand how proteins interact between them, and if they were enriched in specific pathways. STRING analysis found LRRK2 to interact with GRN and GPNMB and is part of a network that also included CD68 and ITGB2 (Fig. 3A). CD68 is known to interact with GRN, CD63, and ORL1, and predicted to co-express with ITGB2 and FCGR1A. Protein enrichment analyses indicate that the 26 proteins are enriched in proteins involved in the leukocyte activation pathway (GO:0045321 leukocyte activation, *P*=0.002) and the microglial cell activation pathway (GO:0001774 microglial cell activation, *P*=0.003), along with enrichment in the lysosome cellular component (GO:0005764 lysosome, *P*=0.001; Fig. 3B). Gene-disease enrichment analyses also identified an enrichment in lysosomal storage disease genes (C0085078 Lysosomal storage disease, *P*=0.005; Fig. 3C). Lysosomal storage disease genes include GRN, GPNMB, GAA, and CHIT1 which, according to STRING analyses, interact with CD68, CD63, and TLR3, and are known to have lysosomal membrane function (Supplementary Fig. 4).

**Figure 3.**
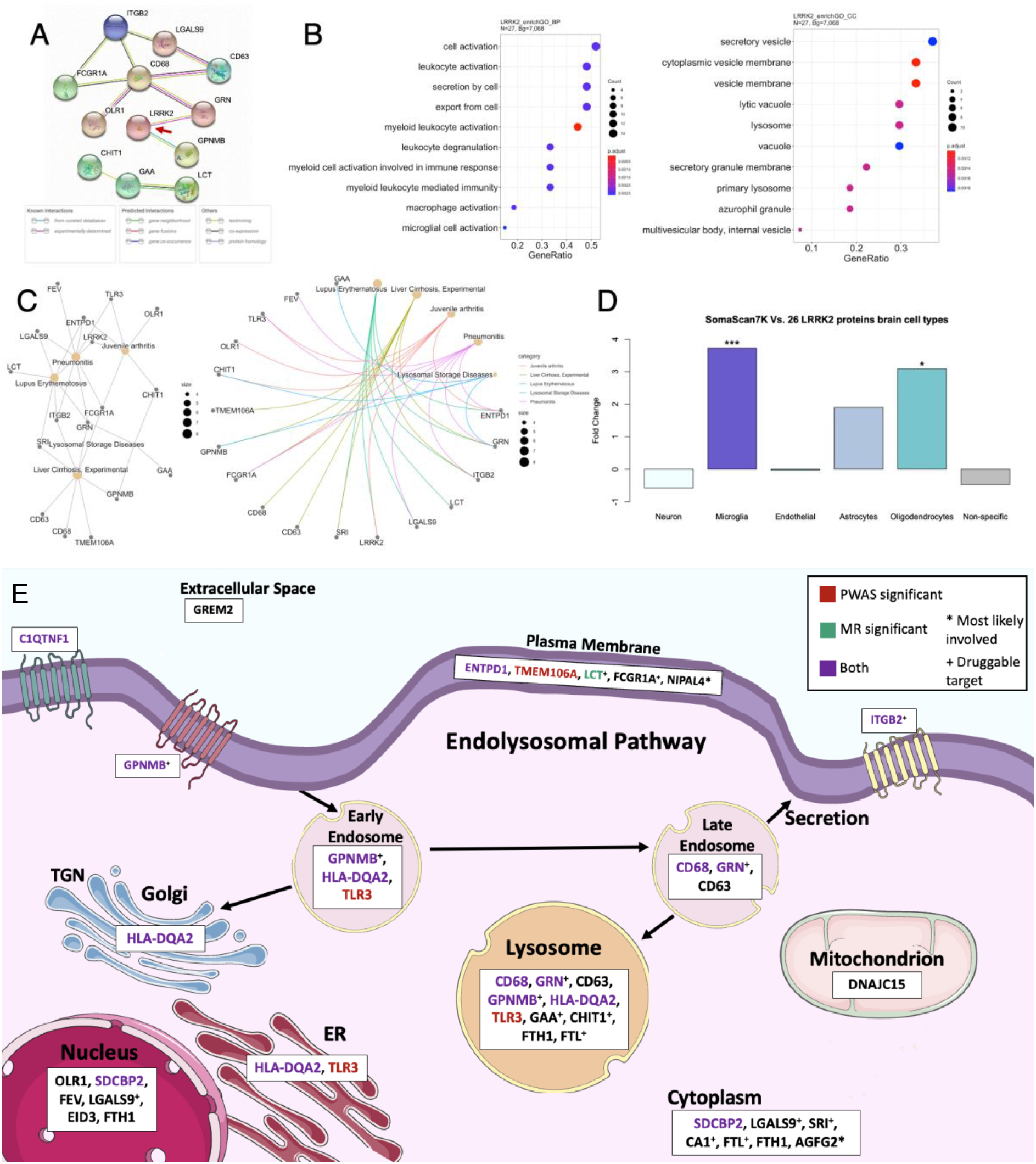
Protein pathways and cell-types. (**A**) STRING interactions of the 26 proteins and LRRK2 (red arrow).(**B**) EnrichGO of 6,138 SomaScan7K genes with recognized unique entrez ID as background and 27 genes of interest (26 proteins and *LRRK2*). Biological process (BP) and cellular component (CC) are shown. (**C**) DisGeNET gene-disease enrichment of 27 genes and 6,138 SomaScan7K genes. The Top 5 disease-gene connections are shown. (**D**) Brain cell-type fold-change (FC) barplot using 5,709 CSF SomaScan7K proteins with brain cell-type data and 26 proteins. *** is FC > |3.7| (*P*=4.91×10^−5^) and * is FC > |3.0|. (**E**) Cellular pathways. Pathway involvement of the 26 LRRK2 associated proteins in the endosome, Golgi apparatus, endoplasmic reticulum (ER), nucleus, mitochondrion, cytoplasm, plasma membrane, extracellular space, and lysosome. A large proportion of LRRK2 associated proteins are involved (or enriched) in the endolysosomal pathway. *Most likely involved. ^+^Druggable protein target. Cell structure images are from Bioicons.

#### Brain Cell-type enrichment Analysis

In line with the gene enrichment analyses, cell-type enrichment analyses indicate enrichment of microglia-specific proteins (Fold change:3.73, *P*=4.91×10^−5^; Fig. 3D; Supplementary Table 7). Nine of the 26 proteins were microglial/macrophage-specific (Supplementary Table 8). Of the eleven PD associated proteins, our analyses indicate that four (CD68, GRN, HLA-DQA2, TLR3) were microglia-specific, SRI was astrocyte-specific, and GPNMB was oligodendrocyte-specific. LRRK2, GRN, and GPNMB had predominant glial (microglia and oligodendrocyte) cell-type expression and were known to interact based on STRING (Fig. 3E).

## Discussion

Genome-wide association studies have identified over 90 loci associated with PD.^8^ However, the genes driving the association for those loci and the functional mechanism by which those signals lead to PD are unknown for most of these loci. In addition, those analyses were not designed to identify complex interactions between proteins. A common approach to identify the functional and causal genes driving the association is QTL mapping combined with colocalization and Mendelian Randomization analyses. This is usually performed with transcriptomic data, but it is now understood that gene-expression levels do not correlate with protein levels.^5^ In fact, proteins are likely closer mechanistically to the disease outcome than gene-expression. A recent study extended the eQTL approach to protein levels by analyzing pQTL for 1,305 proteins and 131 individuals from the PPMI study.^21^ Kaiser et al.^3^ found that pQTLs for GPNMB, HLA-DAQ2, and FCGR2A not only colocalized with PD risk loci but were also significant in the MR analyses, implicating those proteins in the causal pathway of PD. The main limitation of this study is that they only included cis-pQTL, missing on additional causal and novel proteins along with protein-protein interactions.

In recent studies by our group,^4,5^ we used cis and trans-pQTL mapping to identify novel causal and druggable targets for AD and PD. We identified a highly significant trans-pQTL for TREM2 in the *MS4A4A* region that resolved the *MS4A4A* loci for AD risk.^4^ We also generated a multi-tissue (brain, CSF, and plasma) pQTL atlas that was integrated with colocalization, MR, and drug repositioning to identify novel causal proteins and therapies for AD, PD, and other neurodegenerative diseases.^5^ Yang et al.^5^ identified 35 (brain), 13 (CSF), and 15 (plasma) proteins that were PD causal based on MR with more than 50% colocalizing with PD risk loci.

In addition, Yang et al.^5^ identified seven FDA compounds that target those proteins and in the same direction.

Here, we have increased the sample size of our CSF published studies to 3,107 individuals and focused on the *LRRK2* locus to resolve the most significant locus for PD risk and elucidate the downstream mechanism of *LRRK2* common variants. Briefly, we identified 26 proteins associated with PD risk, 15 of them novel. Subsequent TWAS/FUSION-like PWAS approaches determined that ten of those proteins are genetically associated with PD risk and five were causally linked via MR. Among these proteins, GPNMB and HLA-DAQ2 were already identified by Kaiser et al.^3^ in their cis-only pQTL analyses. In addition, we identified other proteins (GRN, CD68, LCT) that are causal based on all our MR analyses including stringent sensitivity analyses. Other proteins (SDCBP2, ENTPD1, ITGB2, and C1QTNF1) that our MR analyses suggest are causal were genetically associated with PD risk based on the PWAS analyses and some (ENTPD1, ITGB2, and C1QTNF1) showed significant differential protein levels in PD cases compared to controls on the PPMI dataset.

The endolysosomal pathway dysfunction is strongly implicated in the pathobiology of PD.^1,3,22^ Endolysosomal system dysfunction cause disruptions in various essential cellular processes such as endocytosis, phagocytosis, macroautophagy, mitophagy, and/or lysosomal function. LRRK2 is a part of endolysosomal pathway and one of its known functions is the involvement in endosome sorting prior to lysosome fusion.^23^ Interestingly, we found that LRRK2 variants modify the protein level of two well-established PD-associated proteins: GRN and GNMPB. GRN localizes in endosomes and lysosomes and GRN protein levels are known to tightly regulate lysosomal function.^24^ In addition to the localization of GPNMB to plasma membrane, it is also present in intracellular vesicles such as endosomes and lysosomes.^25^ Recently, GPNMB was shown to directly interact with α-synuclein, a disease defining protein aggregating in PD.^26^ Membrane protein HLA-DQA2, a protein of HLA family that has been previously implicated in PD pathogenesis,^27^ also localizes in endosomes and lysosomes together with lysosome-associated proteins CD63^28^ and CD68.^29^ Previously, expression of CD68 has been shown to be increased in the p.G2019S LRRK2 transgenic mice compared to control mice upon cerebral injection of α-synuclein pre-formed fibrils.^30^ Importantly, the proteins identified here were not only enriched in the endolysosomal pathway, but other pathways are also related to the intracellular trafficking including the secretory vesicle pathway (GO:0099503 secretory vesicle; *P*=0.002), or the vacuole (GO:0005773 vacuole; *P*=0.002), indicating that changes in protein/vesicle tracking, and not just the lysosome, are important in PD pathogenesis (Figure 3E).

In addition to identifying specific biological pathways, our analyses can also inform about the cell-types in which these pathways are disrupted. We found that the proteins identified in our study were enriched in microglia (Fold change: 3.73; *P*<0.05) and immune response (*P*=0.002). Microglia dysregulation is known to be implicated in AD, but few studies have identified a strong enrichment of microglia genes/proteins and immune function in PD.^1,31^ Indeed, a large proportion of the proteins identified here play a role in immune response. Many of these proteins are localized in plasma membrane of microglia and macrophages (such as ENTPD1 and TLR3), which also localize to lysosomes and FCGR1A, or in extracellular space (e.g., LGALS9 and CHIT1), or both (C1QTNF1, OLR1, ITGB2) (Figure 3E). Intriguingly, many proteins (GPNMB, CD63, and HLA-DQA2) identified to be part of the endolysosomal pathway also localize to the plasma membrane and have a role in immune function.^32-34^ Interestingly, a recent study found that a common non-coding variant of LRRK2 specifically contributes to risk of PD via microglial LRRK2 expression.^35^ Our findings strongly support the previous observations regarding the importance of microglia and immune system function in the underlying pathobiology of PD. Independent of causality, this is the first time that all these proteins have been connected as part of the same pathways, and this helps to understand the mechanism by which *LRRK2* common variants contribute to disease pathogenesis and pathophysiology.

Finally, our study paves the way for clinical trials of new therapeutic approaches. For example, our analyses indicate that GRN levels are part of the causal pathway for PD. Several clinical trials currently target GRN or the GRN receptor (Sortilin) (such as NCT04408625, NCT04747431, and NCT05262023) in *GRN*-mutation carriers, that if effective could be translated to PD. Likewise, the clinical trials targeting ITGB2 (NCT03812263), GAA (NCT00976352), and LCT (NCT02902016, NCT01145586, and NCT05100719) could similarly be translated to PD in the future. Based on the UniProt DrugBank, there are druggable protein targets for CA1 (DB00819), CHIT1 (DB03539), FCGR1A (DB00112), FTL (DB09147), GAA (DB00284), GPNMB (DB05996), LCT (DB04779), LGALS9 (DB04472), and SRI (DB11348). Interestingly, atorvastatin^36^ and celastrol^37^ have been shown to reduce CD68 gene expression, which could similarly be applicable to PD.

In conclusion, trans-pQTL can identify novel protein interactions in an unbiased manner. This study linked, for the first time, LRRK2 variants with proteins that have support in our PWAS, MR, and PPMI differential protein level analyses: some previously implicated in PD (GRN and GPNMB), and others that are novel (C1QTNF1 and ITGB2). Our results suggest that the CSF concentrations of these proteins (such as GRN, GPNMB, C1QTNF1, and ITGB2) have potential as biomarkers for target engagement and disease progression modification in clinical trials that target *LRRK2*.

## Data Availability

Proteomic, pQTL, and raw data from the Knight ADRC participants are available at the NIAGADS and can be accessed at https://www.niagads.org/knight-adrc-collection.
Data from the DIAN cohort is available to qualified investigators and can be requested at https://dian.wustl.edu/our-research/for-investigators/diantu-investigator-resources/dian-tu-biospecimen-request-form/.
ADNI and PPMI proteomic data can be found at: https://adni.loni.usc.edu/ and, https://www.ppmi-info.org/respectively.

https://dian.wustl.edu/our-research/for-investigators/diantu-investigator-resources/dian-tu-biospecimen-request-form/

https://adni.loni.usc.edu/

https://www.ppmi-info.org/

## Abbreviations

ADNI: Alzheimer’s Disease Neuroimaging Initiative;
COJO: Conditional & Joint Association analysis;
CSF: Cerebrospinal fluid;
DIAN: Dominantly Inherited Alzheimer Network;
FDR: False Discovery Rate;
GRN: granulin precursor;
GWAS: Genome-wide association study;
LD: Linkage disequilibrium;
MAP: Memory and Aging Project;
MR: Mendelian randomization;
Pau: Hospital Sant Pau;
PheWAS: Phenome-wide association study;
PPMI: Parkinson’s Progression Markers Initiative;
pQTL: Protein quantitative trail loci;
PWAS: Proteome-wide association studies;
SNP: Single nucleotide polymorphism

## Acknowledgements

We thank all the participants and their families, as well as the many involved institutions and their staff.

This work was supported by access to equipment made possible by the Hope Center for Neurological Disorders, the Neurogenomics and Informatics Center (NGI: https://neurogenomics.wustl.edu/) and the Departments of Neurology and Psychiatry at Washington University School of Medicine.

*ADNI resources:* Data collection and sharing for this project was funded by the Alzheimer’s Disease Neuroimaging Initiative (ADNI) (National Institutes of Health Grant U01 AG024904) and DOD ADNI (Department of Defense award number W81XWH-12-2-0012). ADNI is funded by the National Institute on Aging, the National Institute of Biomedical Imaging and Bioengineering, and through generous contributions from the following: AbbVie, Alzheimer’s Association; Alzheimer’s Drug Discovery Foundation; Araclon Biotech; BioClinica, Inc.; Biogen; Bristol-Myers Squibb Company; CereSpir, Inc.; Cogstate; Eisai Inc.; Elan Pharmaceuticals, Inc.; Eli Lilly and Company; EuroImmun; F. Hoffmann-La Roche Ltd and its affiliated company Genentech, Inc.; Fujirebio; GE Healthcare; IXICO Ltd.; Janssen Alzheimer Immunotherapy Research & Development, LLC.; Johnson & Johnson Pharmaceutical Research & Development LLC.; Lumosity; Lundbeck; Merck & Co., Inc.; Meso Scale Diagnostics, LLC.; NeuroRx Research; Neurotrack Technologies; Novartis Pharmaceuticals Corporation; Pfizer Inc.; Piramal Imaging; Servier; Takeda Pharmaceutica l Company; and Transition Therapeutics. The Canadian Institutes of Health Research is providing funds to support ADNI clinical sites in Canada. Private sector contributions are facilitated by the Foundation for the National Institutes of Health (www.fnih.org). The grantee organization is the Northern California Institute for Research and Education, and the study is coordinated by the Alzheimer’s Therapeutic Research Institute at the University of Southern California. ADNI data are disseminated by the Laboratory for Neuro Imaging at the University of Southern California.

*DIAN resources:* Data collection and sharing for this project was supported by The Dominantly Inherited Alzheimer Network (DIAN, U19AG032438) funded by the National Institute on Aging (NIA),the Alzheimer’s Association (SG-20-690363-DIAN), the German Center for Neurodegenerative Diseases (DZNE), Raul Carrea Institute for Neurological Research (FLENI), Partial support by the Research and Development Grants for Dementia from Japan Agency for Medical Research and Development, AMED, and the Korea Health Technology R&D Project through the Korea Health Industry Development Institute (KHIDI), Spanish Institute of Health Carlos III (ISCIII), Canadian Institutes of Health Research (CIHR), Canadian Consortium of Neurodegeneration and Aging, Brain Canada Foundation, and Fonds de Recherche du Québec – Santé. This manuscript has been reviewed by DIAN Study investigators for scientific content and consistency of data interpretation with previous DIAN Study publications. We acknowledge the altruism of the participants and their families and contributions of the DIAN research and support staff at each of the participating sites for their contributions to this study.

ACE Alzheimer Center Barcelona acknowledges all patients and their families for their collaboration. For CSF biomarker research, A.R. and M.B. received support from the European Union/EFPIA Innovative Medicines Initiative Joint undertaking ADAPTED and MOPEAD projects (grant numbers 115975 and 115985, respectively). M.B. and A.R. are also supported by national grants PI13/02434, PI16/01861, PI17/01474, PI19/01240,PI19/01301, PI22/01403 from the Acción Estratégica en Salud, integrated in the Spanish National RCDCI Plan and funded by Instituto de Salud Carlos III (ISCIII)—Subdirección General de Evaluación and the Fondo Europeo de Desarrollo Regional (FEDER—”Una manera de Hacer Europa”).A.R. and M.B. have also received support from CIBERNED (Instituto de Salud Carlos III (ISCIII). A.R. is also supported by the EXIT project, EU Euronanomed3 Program JCT2017, Grant No. AC17/00100 and PREADAPT project; the Joint Program for Neurodegenerative Diseases (JPND), Grant No. AC19/00097; Acción Estratégica en Salud, integrated in the Spanish National RCDCI Plan and funded by Instituto de Salud Carlos III (ISCIII)— Subdirección General de Evaluación and the Fondo Europeo de Desarrollo Regional (FEDER—”Una manera de Hacer Europa”). I. de Rojas is supported by a national grant from the Instituto de Salud Carlos III FI20/00215.

*Data used in preparation of this article were obtained from the Alzheimer’s Disease Neuroimaging Initiative (ADNI) database (adni.loni.usc.edu). As such, the investigators within the ADNI contributed to the design and implementation of ADNI and/or provided data but did not participate in analysis or writing of this report. A complete listing of ADNI investigators can be found at: http://adni.loni.usc.edu/wp-content/uploads/how_to_apply/ADNI_Acknowledgement_List.pdf

## Supplementary material

Dominantly Inherited Alzheimer Network (DIAN) consortia investigators and coordinators are listed in Supplemental file 2.

