## Supplemental File for "Proteome Wide Association Studies of LRRK2 variants identify novel causal and druggable for Parkinson’s disease"

**Supplementary Table 1 Basic demographics of the cohorts included in the study**
**Supplementary Table 2 Annotation of the variants associated with protein levels on the LRRK2 locus**
**Supplementary Table 3 pQTL SNP p-values**
**Supplementary Table 4 Parkinson's Disease risk table of LRRK2 associated proteins**
**Supplementary Table 5 PWAS p-values and z-scores**
**Supplementary Table 6 PPMI cohort validation**
**Supplementary Table 7 Brain cell types of SomaScan7K and LRRK2 associated proteins**
**Supplementary Table 8 Brain cell types of LRRK2 associated proteins**

**Supplementary references**

**Supplement 2: Dominantly Inherited Alzheimer Network (DIAN) consortia investigators and coordinators**

**Supplementary Figure 1: PheWAS and LD matrix of LRRK2 variants**
**Supplementary Figure 2: Protein-protein correlation**
**Supplementary Figure 3: PPMI significant TWAS Violin plots**
**Supplementary Figure 4: Interaction pathways of LRRK2 associated proteins**

**Supplementary Table 1 Basic demographics of the cohorts included in the study**

| Cohort | # Samples | Avg. Age (SD) | % Male |
|---|---|---|---|
| ADNI | 689 | 73.7 (7.5) | 58.2 |
| DIAN | 193 | 38.6 (10.7) | 48.4 |
| MAP | 805 | 71.4 (8.7) | 46.7 |
| Barcelona-1 | 197 | 68.8 (7.5) | 52.3 |
| Fundació ACE | 438 | 71.9 (8.3) | 41.1 |
| PPMI | 785 | 61.8 (9.4) | 55.1 |

Sample sizes of each of the six cohorts included (3,107 total samples). The three discovery cohorts are DIAN, MAP, and Pau. The three replication cohorts are ADNI, Fundació ACE, and PPMI.
ADNI = Alzheimer's Disease Neuroimaging Initiative; DIAN = Dominantly Inherited Alzheimer Network; MAP = Memory and Aging Project; Barcelona-1 = Hospital Sant Pau; PPMI = Parkinson’s Progression Markers Initiative.

**Supplementary Table 2 Annotation of the variants associated with protein levels on the LRRK2 locus**

| Variant | RSID | Type | Gene : Consequence | MAF | PD SNP $r^2$ |
|---|---|---|---|---|---|
| chr12:10160315:G:A | rs3816844 | Non-coding | *OLR1* : Intron Variant | 0.495 | $1.11\times10^{-4}$ |
| chr12:10178088:G:T | rs113181367 | Non-coding | *TMEM52B* : Intron Variant | 0.075 | $3.35\times10^{-4}$ |
| chr12:33273740:G:A | rs529679490 | N/A | None | 0.006 | $1.57\times10^{-6}$ |
| chr12:39994307:C:T | rs140722239 | Non-coding | *SLC2A13*: Intron Variant | 0.047 | 0.030 |
| chr12:40035894:C:T | rs11564199 | Non-coding | *SLC2A13*: Intron Variant | 0.047 | 0.031 |
| chr12:40039522:G:A | rs78131768 | Non-coding | *SLC2A13* : Intron Variant | 0.047 | 0.031 |
| chr12:40189191:A:G | rs2263418 | Non-coding | *LRRK2-DT* : Non-Coding Transcript Variant | 0.105 | 0.472 |
| chr12:40194013:T:C | rs11175546 | Non-coding | *LRRK2-DT* : Non-Coding Transcript Variant | 0.046 | 0.291 |
| chr12:40198262:T:G | rs79410089 | N/A | None | 0.080 | 0.362 |
| chr12:40204350:T:G | rs11564273 | N/A | None | 0.069 | 0.435 |
| **chr12:40220632:C:T** | **rs76904798** | **Non-coding** | ***LRRK2* : Intron variant** | **0.134** | **1.0** |
| chr12:40259708:G:A | rs28903073 | Non-coding | *LRRK2* : Intron variant | 0.032 | 0.005 |
| chr12:40320097:T:C | rs35303786 | Exonic | *LRRK2* : Missense Variant | 0.021 | 0.003 |
| chr12:40342447:C:T | rs919174 | Non-coding | *LRRK2* : Intron Variant | 0.131 | 0.171 |
| chr12:40397605:G:A | rs117929583 | Non-coding | *MUC19* : Intron Variant | 0.024 | 0.110 |
| chr12:41349642:C:T | rs190000583 | Non-coding | *PDZRN4* : Intron Variant | 0.036 | 0.003 |

Variant name, Rs-id, and intron/exon type of the 16 independently associated chromosome 12 SNPs ordered by bp position. Only rs35303786 (chr12:40320097:T:C) is known to be exonic. Top PD and CSF *GRN* SNP rs76904798 (chr12:40220632:C:T) is shown in bold.

**Supplementary Table 3 pQTL SNP P-values**

| Protein | Aptamer | SNP #1 | SNP #2 | SNP #3 | SNP #4 | SNP #5 |
|---|---|---|---|---|---|---|
| GPNMB | X8240.207 | $4.79\times10^{-304}$ | $3.16\times10^{-28}$ | $4.79\times10^{-8}$ | | |
| GPNMB | X5080.131 | $6.76\times10^{-225}$ | $1.45\times10^{-33}$ | $7.41\times10^{-9}$ | | |
| CHIT1 | X3600.2 | $3.47\times10^{-175}$ | $3.63\times10^{-11}$ | $1.00\times10^{-8}$ | $2.24\times10^{-8}$ | |
| LCT | X9017.58 | $5.89\times10^{-142}$ | $3.47\times10^{-8}$ | $3.72\times10^{-8}$ | | |
| TLR3 | X16918.198 | $4.68\times10^{-137}$ | $2.09\times10^{-12}$ | | | |
| HLA-DQA2 | X7757.5 | $1.07\times10^{-116}$ | $1.38\times10^{-28}$ | $5.01\times10^{-12}$ | $6.92\times10^{-9}$ | $7.08\times10^{-9}$ |
| GPNMB | X8606.39 | $2.75\times10^{-103}$ | | | | |
| CD68 | X20528.23 | $4.37\times10^{-63}$ | $1.29\times10^{-10}$ | $3.72\times10^{-9}$ | $5.37\times10^{-9}$ | |
| CD68 | X18922.27 | $1.58\times10^{-57}$ | $4.37\times10^{-10}$ | $2.34\times10^{-9}$ | $1.91\times10^{-8}$ | |
| C1QTNF1 | X6304.8 | $1.86\times10^{-30}$ | $1.41\times10^{-9}$ | | | |
| ENTPD1 | X3182.38 | $3.72\times10^{-29}$ | $7.59\times10^{-9}$ | | | |
| LGALS9 | X9197.4 | $1.45\times10^{-27}$ | $3.09\times10^{-12}$ | | | |
| GRN | X4992.49 | $1.26\times10^{-20}$ | $3.16\times10^{-17}$ | | | |
| AGFG2 | X23597.11 | $8.71\times10^{-20}$ | $3.39\times10^{-9}$ | | | |
| ITGB2 | X12750.9 | $1.91\times10^{-17}$ | $2.82\times10^{-15}$ | $4.27\times10^{-8}$ | | |
| TMEM106A | X10499.1 | $2.57\times10^{-17}$ | $7.59\times10^{-10}$ | | | |
| OLR1 | X3636.37 | $6.03\times10^{-16}$ | $1.23\times10^{-10}$ | | | |
| OLR1 | X7893.19 | $4.79\times10^{-15}$ | $4.07\times10^{-10}$ | | | |
| GAA | X9385.4 | $2.24\times10^{-10}$ | | | | |
| SDCBP2 | X19261.12 | $5.37\times10^{-10}$ | | | | |
| GREM2 | X5598.3 | $7.94\times10^{-9}$ | $3.98\times10^{-8}$ | | | |
| NIPAL4 | X12864.9 | $4.79\times10^{-8}$ | | | | |
| CA1 | X4969.2 | $7.24\times10^{-8}$ | | | | |
| FCGR1A | X3312.64 | $8.51\times10^{-8}$ | | | | |
| SRI | X12356.65 | $1.10\times10^{-7}$ | | | | |
| FTL | X15324.58 | $1.48\times10^{-7}$ | | | | |
| FEV | X12740.55 | $1.62\times10^{-7}$ | | | | |
| EID3 | X8079.39 | $1.66\times10^{-7}$ | | | | |
| FTL | X5934.1 | $3.80\times10^{-7}$ | | | | |
| CD63 | X9190.7 | $5.75\times10^{-7}$ | | | | |
| DNAJC15 | X7197.2 | $6.46\times10^{-7}$ | | | | |

SNP p-values of the top SNPs for the 31 significant aptamers. Sorted by SNP #1 p-value.

**Supplementary Table 4 Parkinson's disease risk table of LRRK2 associated proteins**

| Gene | PD | Neurodegeneration | Function/Expression | Experiment Type | Citation |
|---|---|---|---|---|---|
| ***CD63*** | **Yes** | Yes | Decreased in PD | Multiplex immunoassay of circulating small EVs (sEVs) | [Picca et al., 2020] |
| ***ENTPD1*** | **Yes** | Yes | Decreased in PD | RT-qPCR of substantia nigra mRNA | [Garcia-Esparcia et al., 2015] |
| ***GRN*** | **Yes** | Yes | Decreased in PD | Meta-analysis of PD GWAS | [Nalls et al., 2019] |
| ***HLA-DQA2*** | **Yes** | Yes | HLA region PD association | Haplotype analysis and step-wise conditional analysis | [Hill-Burns et al., 2011] |
| ***GAA*** | **Yes** | Yes | Increased in PD | Lysosomal enzymatic activity assay | [Alcalay et al., 2018] |
| ***GPNMB*** | **Yes** | Yes | Increased in sporadic PD | Induction of lipidopathy and immunohistochemical staining | [Moloney et al., 2018] |
| ***SRI*** | **Yes** | Yes | Increased in PD | Immunoprecipitation and cell lines | [Genovese et al., 2020] |
| ***LCT*** | **Yes** | Yes | Increased in PD | Two-sample Mendelian Randomization | [Domenighetti et al, 2022] |
| ***CD68*** | **Yes** | Yes | *LRRK2* Expression | Human and mouse cell culture Immunohistochemistry | [Xu et al., 2020] Preprint |
| ***EID3*** | **Yes** | Yes | PD candidate gene | Differential expression analysis and PPI networks | [George et al., 2019] |
| ***TLR3*** | **Yes** | Yes | Reduced risk EOPD | SNP genotyping and odds ratios analysis | [Wang et al., 2020] |
| *C1QTNF1* | Related | Yes | Downregulated in PD patients | PD-patient specific dopaminergic cultures DE analysis | [Momcilovic et al., 2016] |
| *DNAJC15* | Related | Yes | OGC pesticide exposure PD DML | Genome-scale methylation profiling | [Go et al., 2020] |
| *AGFG2* | No | Yes | AD pathology | Differential gene expression (DGE) analyses | [Fernandez et al., 2022] |
| *ITGB2* | No | Yes | Aging and neurodegeneration | Human aging and neurodegenerative disease microarray datasets | [Mukherjee et al., 2019] |
| *OLR1* | No | Yes | Neuroinflammatory gene | Single-nuclei sequencing | [Agarwal et al., 2020] |
| *SDCBP2* | No | Yes | Neurexin protein interactor | Protein–protein interaction network and co-expression analysis | [Cuttler et al., 2021] |
| *GREM2* | No | Yes | Neuroprotection by downregulating *GREM2* genes | Microarray expression profiling | [Forcella et al., 2020] |
| *CHIT1* | No | Yes | Microglia/macrophage activation in ALS | LC-MS/MS CSF proteomics | [Karayel et al., 2022] |
| *FTL* | No | Yes | Neurodegeneration, brain iron accumulation | Postmortem brain iron histochemistry, mtDNA variant analysis | [Kurzawa-Akanbi et al., 2021] |
| *FEV* | No | Related | Serotonergic (5-HT) neuron expression | Whole-exome data analyses and Biallelic burden calculations | [Doan et al, 2019] |
| *LGALS9* | No | Related | Enhances microglial TNF production | Glial culture immunocytochemistry and cytokine measurement | [Steelman et al., 2014] |
| *CA1* | No | Related | Neuropathic pain | Carbonic anhydrase inhibition and Ischaemic brain damage | [Dettori et al., 2021] |
| *TMEM106A* | No | No | Increased in AD, *TMEM106B* paralog | qPCR, western blot, and immunohistochemistry | [Zhao et al., 2021] |
| *NIPAL4* | No | No | Congenital Ichthyosiform Erythroderma | Mutational screening and sequence pathogenicity prediction | [Laadhar et al., 2020] |
| *FCGR1A* | No | No | Leptomeningeal Metastasis Biomarker | LC-MS/MS CSF proteomics and protein microarray analysis | [Juanes-Velasco et al., 2022] |

Proteins are organized by having prior studies involving PD risk and/or neurodegeneration, such as AD and ALS, and neurological disorders such as stroke. The columns are the protein name, whether the protein has been shown to be associated with PD, whether there has been an article on the protein being involved in neurodegeneration, the function/effect of the protein in the chosen article, types of experiments done in the article to prove association, and the article's citation. The 11 proteins with predicted PD risk gene association are shown in bold.

**Supplementary Table 5 PWAS p-values and z-scores**

| Protein | Aptamer | PWAS.P-value | PWAS.Z |
|---|---|---|---|
| HLA-DQA2 | X7757.5 | **$2.61\times10^{-47}$** | 14.44715 |
| ITGB2 | X12750.9 | **$4.49\times10^{-47}$** | 14.40981 |
| C1QTNF1 | X6304.8 | **$6.96\times10^{-46}$** | 14.21923 |
| GRN | X4992.49 | **$4.32\times10^{-42}$** | 13.59439 |
| GPNMB | X8240.207 | **$2.32\times10^{-41}$** | 13.47087 |
| GPNMB | X5080.131 | **$1.48\times10^{-38}$** | 12.98536 |
| ENTPD1 | X3182.38 | **$1.59\times10^{-34}$** | 12.25466 |
| TMEM106A | X10499.1 | **$1.51\times10^{-32}$** | 11.87987 |
| CD68 | X20528.23 | **$3.66\times10^{-25}$** | 10.36289 |
| CD68 | X18922.27 | **$6.83\times10^{-24}$** | 10.07916 |
| SDCBP2 | X19261.12 | **$2.88\times10^{-18}$** | 8.71598 |
| TLR3 | X16918.198 | **$1.35\times10^{-15}$** | 7.98963 |
| OLR1 | X3636.37 | 0.576 | -0.55992 |
| OLR1 | X7893.19 | 0.717 | -0.36238 |
| AGFG2 | X23597.11 | NA | NA |
| CA1 | X4969.2 | NA | NA |
| CD63 | X9190.7 | NA | NA |
| CHIT1 | X3600.2 | NA | NA |
| DNAJC15 | X7197.2 | NA | NA |
| EID3 | X8079.39 | NA | NA |
| FCGR1A | X3312.64 | NA | NA |
| FEV | X12740.55 | NA | NA |
| FTL | X5934.1 | NA | NA |
| FTL | X15324.58 | NA | NA |
| GAA | X9385.4 | NA | NA |
| GPNMB | X8606.39 | NA | NA |
| GREM2 | X5598.3 | NA | NA |
| LCT | X9017.58 | NA | NA |
| LGALS9 | X9197.4 | NA | NA |
| NIPAL4 | X12864.9 | NA | NA |
| SRI | X12356.65 | NA | NA |

PWAS top hits from TWAS/FUSION. 12 aptamers (10 proteins) (bolded) had significant PWAS p-values. Sorted by PWAS p-values. PWAS p-value as NA refers to TWAS/FUSION excluding the gene from the analysis due to the aptamer's SNP-heritability p-value < 0.05.

**Supplementary Table 6 PPMI cohort validation**

| Protein | FDR SNP Direction | PWAS p-value | PPMI Direction | Control Vs. Case | Control Vs. Prodromal | Control Vs. LRRK2+ | Control Vs. GBA+ | Control Vs. SNCA+ |
|---|---|---|---|---|---|---|---|---|
| **HLA-DQA2** | ++ | $2.61\times10^{-47}$ | ++ | * | *** | *** | NS | NS |
| **ITGB2** | ++ | $4.49\times10^{-47}$ | ++ | NS | *** | *** | NS | NS |
| **C1QTNF1** | ++ | $6.96\times10^{-46}$ | ++ | NS | *** | *** | NS | NS |
| **GRN** | ++ | $4.32\times10^{-42}$ | ++ | NS | *** | ** | NS | NS |
| **GPNMB (X8240.207)** | ++ | $2.32\times10^{-41}$ | ++ | NS | *** | *** | NS | NS |
| **GPNMB (X5080.131)** | ++ | $1.48\times10^{-38}$ | ++ | NS | *** | *** | NS | NS |
| **ENTPD1** | ++ | $1.59\times10^{-34}$ | ++ | NS | ** | ** | NS | NS |
| TMEM106A | ++ | $1.51\times10^{-32}$ | ++ | NS | *** | NS | NS | NS |
| OLR1 (X3636.37) | -- | 0.576 | NS | NS | * | NS | NS | NS |
| OLR1 (X7893.19) | -- | 0.717 | NS | NS | * | NS | NS | NS |
| **FEV** | -- | NA | NS | * | NS | NS | NS | NS |
| **GREM2** | ++ | NA | ++ | ** | ** | NS | NS | NS |
| **GAA** | ++ | NA | -- | *** | * | NS | NS | NS |
| **LCT** | -- | NA | -- | NS | ** | ** | * | NS |
| **LGALS9** | ++ | NA | ++ | NS | ** | *** | NS | NS |
| DNAJC15 | -- | NA | NS | NS | NS | NS | ** | NS |
| CA1 | -- | NA | NS | NS | NS | NS | NS | NS |
| CD63 | ++ | NA | NS | NS | NS | NS | NS | NS |
| CHIT1 | -- | NA | NS | NS | NS | NS | NS | NS |
| EID3 | -- | NA | NS | NS | NS | NS | NS | NS |
| FCGR1A | ++ | NA | NS | NS | NS | NS | NS | NS |
| FTL (X5934.1) | -- | NA | NS | NS | NS | NS | NS | NS |
| GPNMB (X8606.39) | ++ | NA | NS | NS | NS | NS | NS | NS |
| NIPAL4 | ++ | NA | NS | NS | NS | NS | NS | NS |
| SRI | -- | NA | NS | NS | NS | NS | NS | NS |

Direction of the top FDR corrected SNP, PWAS p-value, direction of PPMI protein levels, and violin plot significance of control vs. case, prodromal, and mutation (*LRRK2*+, *GBA*+, *SNCA*+) of the 22 proteins (25 aptamers) with PPMI cohort protein data. Proteins are sorted by PWAS p-value. Bolded proteins are control vs. *LRRK2*+ and/or control vs. case significant. Only HLA-DQA2 is significant for both case Vs. control and case Vs. *LRRK2*+. PWAS p-value as NA refers to TWAS/FUSION excluding the gene from the analysis due to the aptamer's SNP-heritability p-value < 0.05.
NS (not significant).

**Supplementary Table 7 Brain cell types of SomaScan7K and LRRK2 locus associated proteins**

| Brain Cell Type | SomaScan7K Proteins (>50% total expression) | 26 Proteins (>50% total expression) | Enrichment (FC / p-value) |
|---|---|---|---|
| Neuron | 525 (9.20%) | 1 (3.85%) | -0.58 / 0.241 |
| Microglia/Macrophage | 418 (7.32%) | 9 (34.61%) | **3.73 / $4.91\times10^{-5}$** |
| Endothelial | 227 (3.98%) | 1 (3.85%) | -0.03 / 0.376 |
| Mature Astrocyte | 227 (3.98%) | 3 (11.54%) | 1.90 / 0.064 |
| Oligodendrocyte | 108 (1.89%) | 2 (7.69%) | 3.09 / 0.073 |
| Non-specific | 4204 (73.64%) | 10 (38.46%) | -0.47 / **$1.31\times10^{-4}$** |

Unique proteins from CSF SomaScan7K with cell type data (N=5,709) and 26 LRRK2 associated proteins. The most common cell-type specific (>50% total expression) cell type in the CSF SomaScan7K was neuronal. The most common cell type of the 26 LRRK2 associated proteins was microglial/macrophage. Enrichment p-values by hypergeometric distribution using dhyper function in R.

**Supplementary Table 8 Brain cell types of LRRK2 associated proteins**

| Gene | Max Cell Proportion | Max Cell Type |
|---|---|---|
| *C1QTNF1* | 0.71 | Endothelial |
| *AGFG2* | 0.66 | Mature astrocyte |
| *SRI* | 0.59 | Mature astrocyte |
| *SDCBP2* | 0.58 | Mature astrocyte |
| *OLR1* | 0.93 | Microglia/Macrophage |
| *ITGB2* | 0.91 | Microglia/Macrophage |
| *CD68* | 0.91 | Microglia/Macrophage |
| *FCGR1A* | 0.87 | Microglia/Macrophage |
| *TMEM106A* | 0.85 | Microglia/Macrophage |
| *HLA-DQA2* | 0.83 | Microglia/Macrophage |
| *TLR3* | 0.79 | Microglia/Macrophage |
| *LGALS9* | 0.60 | Microglia/Macrophage |
| *GRN* | 0.56 | Microglia/Macrophage |
| *ENTPD1* | 0.49 | Microglia/Macrophage |
| *FTL* | 0.45 | Microglia/Macrophage |
| *GAA* | 0.42 | Microglia/Macrophage |
| *EID3* | 0.42 | Microglia/Macrophage |
| *GREM2* | 0.84 | Neuron |
| *NIPAL4* | 0.82 | Oligodendrocyte |
| *GPNMB* | 0.68 | Oligodendrocyte |
| *CD63* | 0.37 | Mixed |
| *DNAJC15* | 0.31 | Mixed |
| *CA1* | 0.22 | Mixed |
| *FEV* | 0.20 | Mixed |
| *LCT* | 0.20 | Mixed |
| *CHIT1* | 0.20 | Mixed |

The sum column is the sum of the proportions of human mature astrocytes, neurons, microglia/macrophages, oligodendrocytes, and endothelial expression. Mixed cell type refers to either no cell type had a max proportion of expression (>40%) or all cell types had equal proportions. Genes are sorted by cell type and max cell proportion.

**Supplement 2: Dominantly Inherited Alzheimer Network (DIAN) consortia investigators and coordinators**

| Name | Email | Affiliation |
|---|---|---|
| Sarah Adams | | Washington University in St. Louis School of Medicine |
| Ricardo Allegri | | Institute of Neurological Research Fleni, Buenos Aires, Argentina |
| Aki Araki | | Niigata University |
| Nicolas Barthelemy | | Washington University in St. Louis School of Medicine |
| Randall Bateman | | Washington University in St. Louis School of Medicine |
| Jacob Bechara | | Neuroscience Research Australia |
| Tammie Benzinger | | Washington University in St. Louis School of Medicine |
| Sarah Berman | | University of Pittsburgh |
| Courtney Bodge | | Brown University-Butler Hospital |
| Susan Brandon | | Washington University in St. Louis School of Medicine |
| William (Bill) Brooks | | Neuroscience Research Australia |
| Jared Brosch | | Indiana University |
| Jill Buck | | Indiana University |
| Virginia Buckles | | Washington University in St. Louis School of Medicine |
| Kathleen Carter | | Emory University School of Medicine |
| Lisa Cash | | Washington University in St. Louis School of Medicine |
| Charlie Chen | | Washington University in St. Louis School of Medicine |
| Jasmeer Chhatwal | | Brigham and Women's Hospital–Massachusetts General Hospital |
| Patricio Chrem | | Institute of Neurological Research Fleni, Buenos Aires, Argentina |
| Jasmin Chua | | Washington University in St. Louis School of Medicine |
| Helena Chui | | University of Southern California |
| Carlos Cruchaga | | Washington University in St. Louis School of Medicine |
| Gregory S Day | | Mayo Clinic Jacksonville |
| Chrismary De La Cruz | | Columbia University |
| Darcy Denner | | Washington University in St. Louis School of Medicine |
| Anna Diffenbacher | | German Center for Neurodegnerative Diseases (DZNE) Munich |
| Aylin Dincer | | Washington University in St. Louis School of Medicine |
| Tamara Donahue | | Washington University in St. Louis School of Medicine |
| Jane Douglas | | University College London |
| Duc Duong | | Emory University School of Medicine |
| Noelia Egido | | Institute of Neurological Research Fleni, Buenos Aires, Argentina |
| Bianca Esposito | | Icahn School of Medicine at Mount Sinai |
| Anne Fagan | | Washington University in St. Louis School of Medicine |
| Marty Farlow | | Indiana University |
| Becca Feldman | | Washington University in St. Louis School of Medicine |
| Colleen Fitzpatrick | | Brigham and Women's Hospital-Massachusetts |
| Shaney Flores | | Washington University in St. Louis School of Medicine |
| Nick Fox | | University College London |
| Erin Franklin | | Washington University in St. Louis School of Medicine |
| Nelly Friedrichsen | | Washington University in St. Louis School of Medicine |
| Hisako Fujii | | Osaka City University |

| | | |
|---|---|---|
| Samantha Gardener | | Edith Cowan University, Perth |
| Bernardino Ghetti | | Indiana University |
| Alison Goate | | Icahn School of Medicine at Mount Sinai |
| Sarah Goldberg | | University of Pittsburgh |
| Jill Goldman | | Columbia University |
| Alyssa Gonzalez | | Washington University in St. Louis School of Medicine |
| Brian Gordon | | Washington University in St. Louis School of Medicine |
| Susanne Gräber-Sultan | @dzne.de | DZNE-Tübingen |
| Neill Graff-Radford | | Mayo Clinic Jacksonville |
| Morgan Graham | | Mayo Clinic Jacksonville |
| Julia Gray | | Washington University in St. Louis School of Medicine |
| Emily Gremminger | | Washington University in St. Louis School of Medicine |
| Miguel Grilo | | University College London |
| Alex Groves | | Washington University in St. Louis School of Medicine |
| Christian Haass | | Ludwig-Maximilians University - Munich |
| Lisa Häsler | | German Center for Neurodegenerative Diseases (DZNE), Tübingen |
| Jason Hassenstab | | Washington University in St. Louis School of Medicine |
| Cortaiga Hellm | | Washington University in St. Louis School of Medicine |
| Elizabeth Herries | | Washington University in St. Louis School of Medicine |
| Laura Hoechst-Swisher | | Washington University in St. Louis School of Medicine |
| Anna Hofmann | | German Center for Neurodegenerative Diseases (DZNE), Tübingen |
| David Holtzman | | Washington University in St. Louis School of Medicine |
| Russ Hornbeck | | Washington University in St. Louis School of Medicine |
| Yakushev Igor | | German Center for Neurodegnerative Diseases (DZNE) Munich |
| Ryoko Ihara | | Tokyo University |
| Takeshi Ikeuchi | | Niigata University |
| Snezana Ikonomovic | | University of Pittsburgh |
| Kenji Ishii | | Niigata University/Tokyo University |
| Clifford Jack | | Mayo Clinic Rochester |
| Gina Jerome | | Washington University in St. Louis School of Medicine |
| Erik Johnson | | Emory University School of Medicine |
| Mathias Jucker | | German Center for Neurodegenerative Diseases (DZNE), Tübingen |
| Celeste Karch | | Washington University in St. Louis School of Medicine |
| Stephan Käser | | German Center for Neurodegenerative Diseases (DZNE), Tübingen |
| Kensaku Kasuga | | Niigata University |
| Sarah Keefe | | Washington University in St. Louis School of Medicine |
| William (Bill) Klunk | | University of Pittsburgh |
| Robert Koeppe | | University of Michigan |
| Deb Koudelis | | Washington University in St. Louis School of Medicine |
| Elke Kuder-Buletta | | German Center for Neurodegenerative Diseases (DZNE), Tübingen |
| Christoph Laske | | German Center for Neurodegenerative Diseases (DZNE), Tübingen |
| Allan Levey | | Emory University School of Medicine |

| | | |
|---|---|---|
| Johannes Levin | | German Center for Neurodegnerative Diseases (DZNE) Munich |
| Yan Li | | Washington University in St. Louis School of Medicine |
| Oscar Lopez | | University of Pittsburgh |
| Jacob Marsh | | Washington University in St. Louis School of Medicine |
| Rita Martinez | | Washington University in St. Louis School of Medicine |
| Ralph Martins | | Edith Cowan University |
| Neal Scott Mason | | University of Pittsburgh Medical Center |
| Colin Masters | | University of Melbourne |
| Kwasi Mawuenyega | | Washington University in St. Louis School of Medicine |
| Austin McCullough | | Washington University in St. Louis School of Medicine |
| Eric McDade | | Washington University in St. Louis School of Medicine |
| Arlene Mejia | | Columbia University |
| Estrella Morenas-Rodriguez | Estrella.Morenas-Rodriguez@dzne.d | Ludwig-Maximilians University, Munich |
| John Morris | | Washington University in St. Louis School of Medicine |
| James MountzMD | | University of Pittsburgh |
| Cath Mummery | | University College London |
| Neelesh Nadkarni | | University of Pittsburgh |
| Akemi Nagamatsu | mail: | Tokyo University |
| Katie Neimeyer | | Columbia University |
| Yoshiki Niimi | | Tokyo University |
| James Noble | | Columbia University |
| Joanne Norton | | Washington University in St. Louis School of Medicine |
| Brigitte Nuscher | | Ludwig-Maximilians University, Munich |
| Antoinette O'Connor | antoinette.o' | University College London |
| Ulricke Obermüller | | Hertie Institute for Clinical Brain Research |
| Riddhi Patira | | University of Pittsburgh |
| Richard Perrin | | Washington University in St. Louis School of Medicine |
| Lingyan Ping | | Emory University School of Medicine |
| Oliver Preische | | German Center for Neurodegenerative Diseases (DZNE), Tübingen |
| Alan Renton | | Icahn School of Medicine at Mount Sinai |
| John Ringman | | University of Southern California |
| Stephen Salloway | | Brown University-Butler Hospital |
| Peter Schofield | | Neuroscience Research Australia |
| Michio Senda | | Osaka City University |
| Nick Seyfried | | Emory University School of Medicine |
| Kristine Shady | | Washington University in St. Louis School of Medicine |
| Hiroyuki Shimada | | Osaka City University |
| Wendy Sigurdson | | Washington University in St. Louis School of Medicine |
| Lori Smith | | University of Pittsburgh |
| Jennifer Smith | | Washington University in St. Louis School of Medicine |
| Beth Snitz | | University of Pittsburgh |
| Hamid Sohrabi | | Edith Cowan University |
| Sochenda Stephens | | Mayo Clinic Jacksonville |

| | | |
|---|---|---|
| Kevin Taddei | | Edith Cowan University |
| Sarah Thompson | | University of Pittsburgh |
| Jonathan Vöglein | | German Center for Neurodegnerative Diseases (DZNE) Munich |
| Peter Wang | | Washington University in St. Louis School of Medicine |
| Qing Wang | | Washington University in St. Louis School of Medicine |
| Elise Weamer | | University of Pittsburgh |
| Chengjie Xiong | | Washington University in St. Louis School of Medicine |
| Jinbin Xu | | Washington University in St. Louis School of Medicine |
| Xiong Xu | | Washington University in St. Louis School of Medicine |

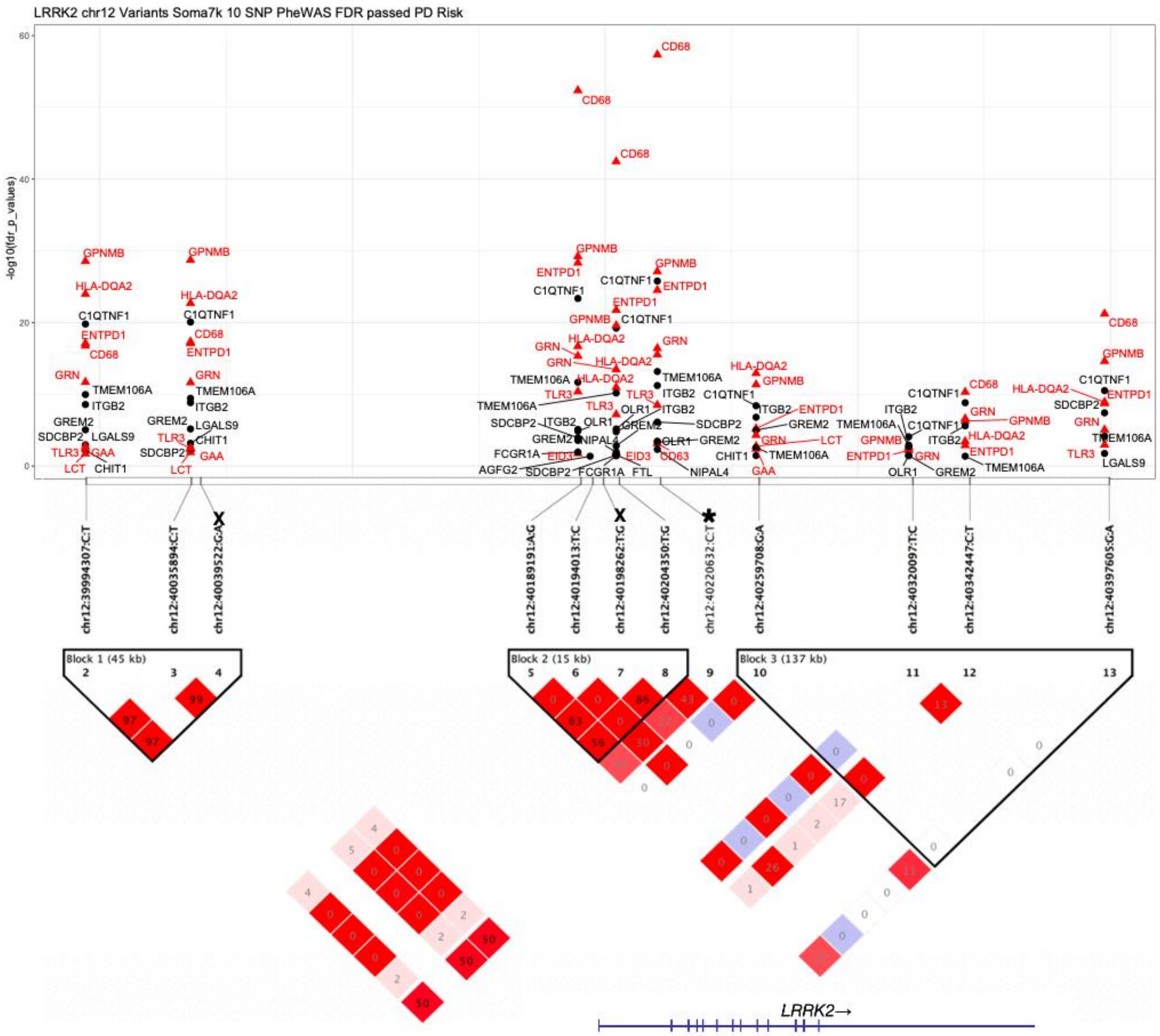


**Supplementary Figure 1: PheWAS and LD matrix of *LRRK2* variants.** PheWAS plot of 10 SNPs in chr12 LRRK2. FDR corrected -log10(p-value) y-axis. 26 unique CSF proteins pass FDR with 11 having known PD risk association (red triangles). LD plot of 12 independently associated Chr12 SNPS, 10 are present with chr12:40220632:C:T (*) and $r^2$ > 0.85 per LD block removed (X). The 5 independently associated SNPs within the *LRRK2* region (Chr12:40,196,744-40,369,285) is below the LD plot with a blue line.

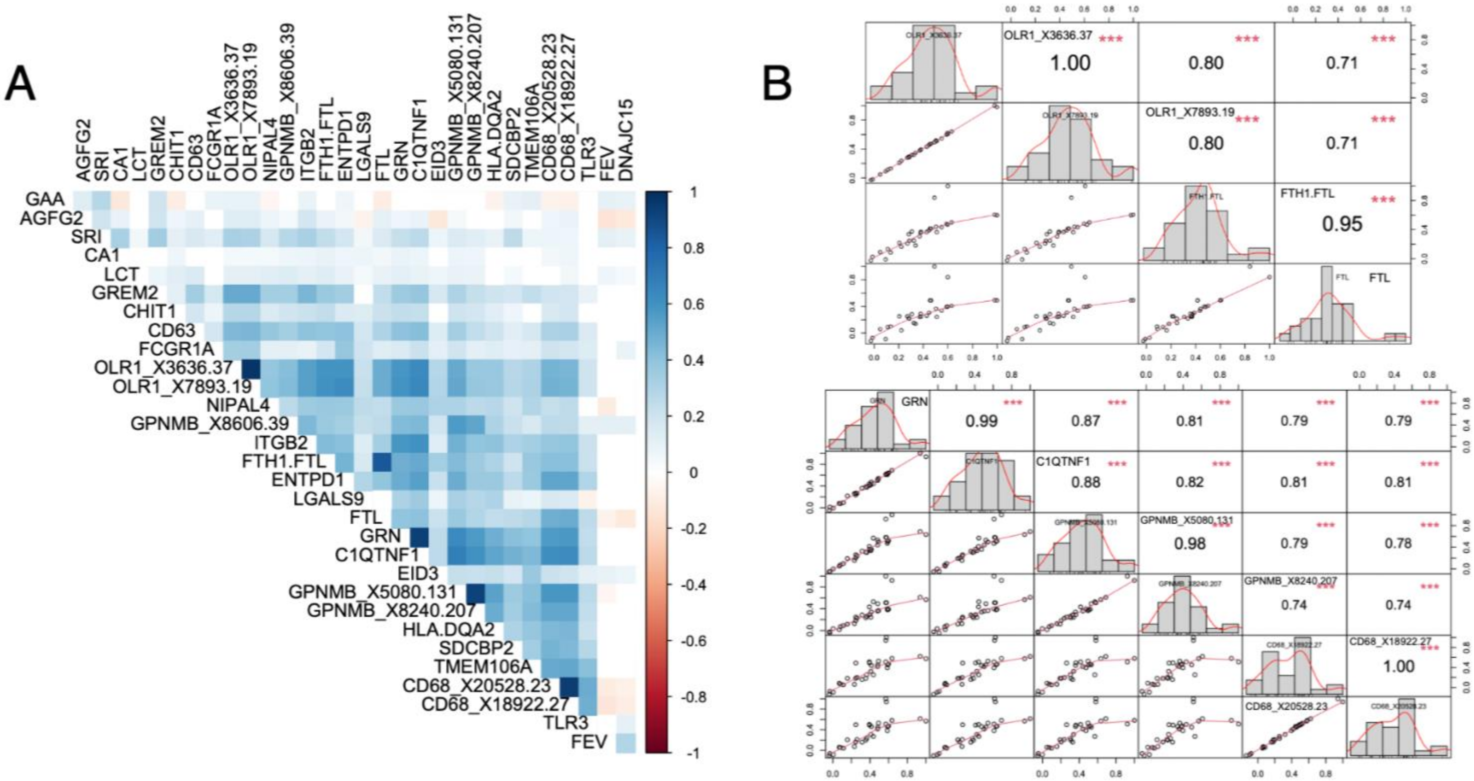


**Supplementary Figure 2: Protein-protein correlation.** (A) Pearson correlation coefficient matrix of the 26 proteins. (B) Chart correlation of the significant regions of OLR1 and FTL analytes above and GRN below. The distribution of each protein is shown on the diagonal with the bottom displaying the bivariate scatterplots with a fitted line. The top of the diagonal shows the correlation value and the significance level as stars with p-value equal to or less than 0.001 (***), 0.01 (**), and 0.05 (*).

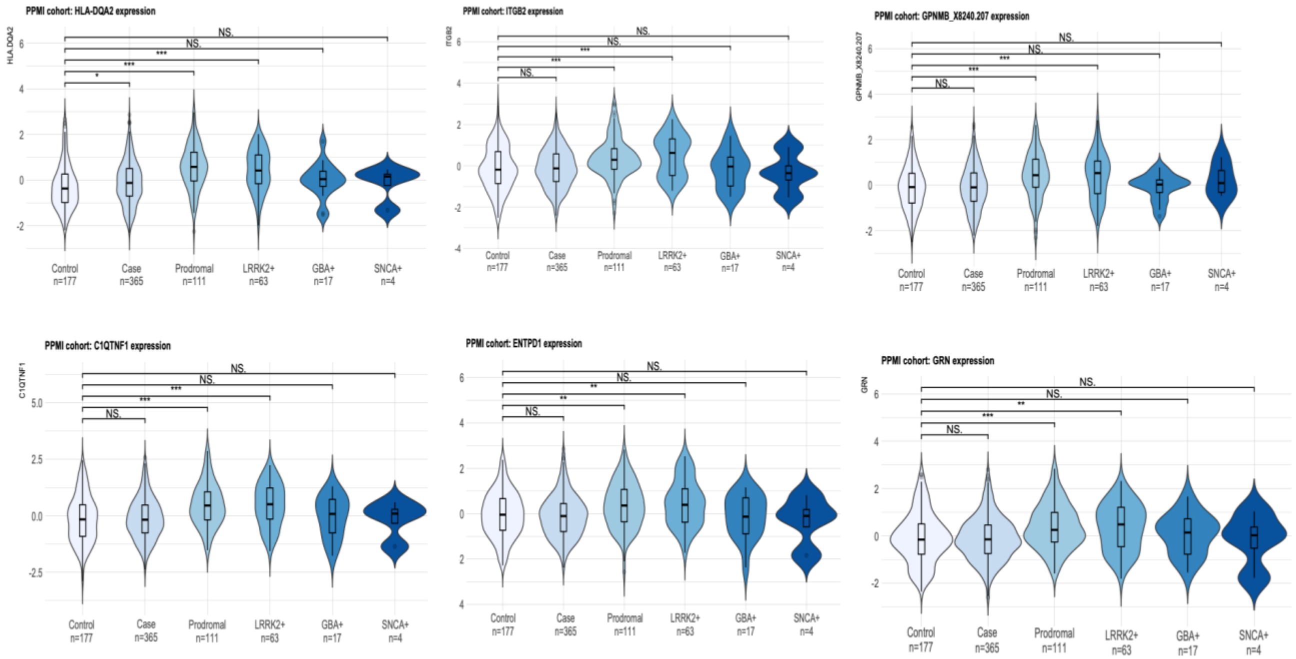


**Supplementary Figure 3: PPMI significant PWAS Violin plots.** Plot of *HLA-DQA2*, *C1QTNF1*, *ITGB2*, *ENTPD1*, *GPNMB*, & *GRN* gene expression. Control Vs. PD case and mutation carriers (*LRRK2*$^{+}$, *GBA*$^{+}$, and *SNCA*$^{+}$) (light to dark blue). The significance level as stars with p-value equal to or less than 0.001 (***), 0.01 (**), and 0.05 (*).

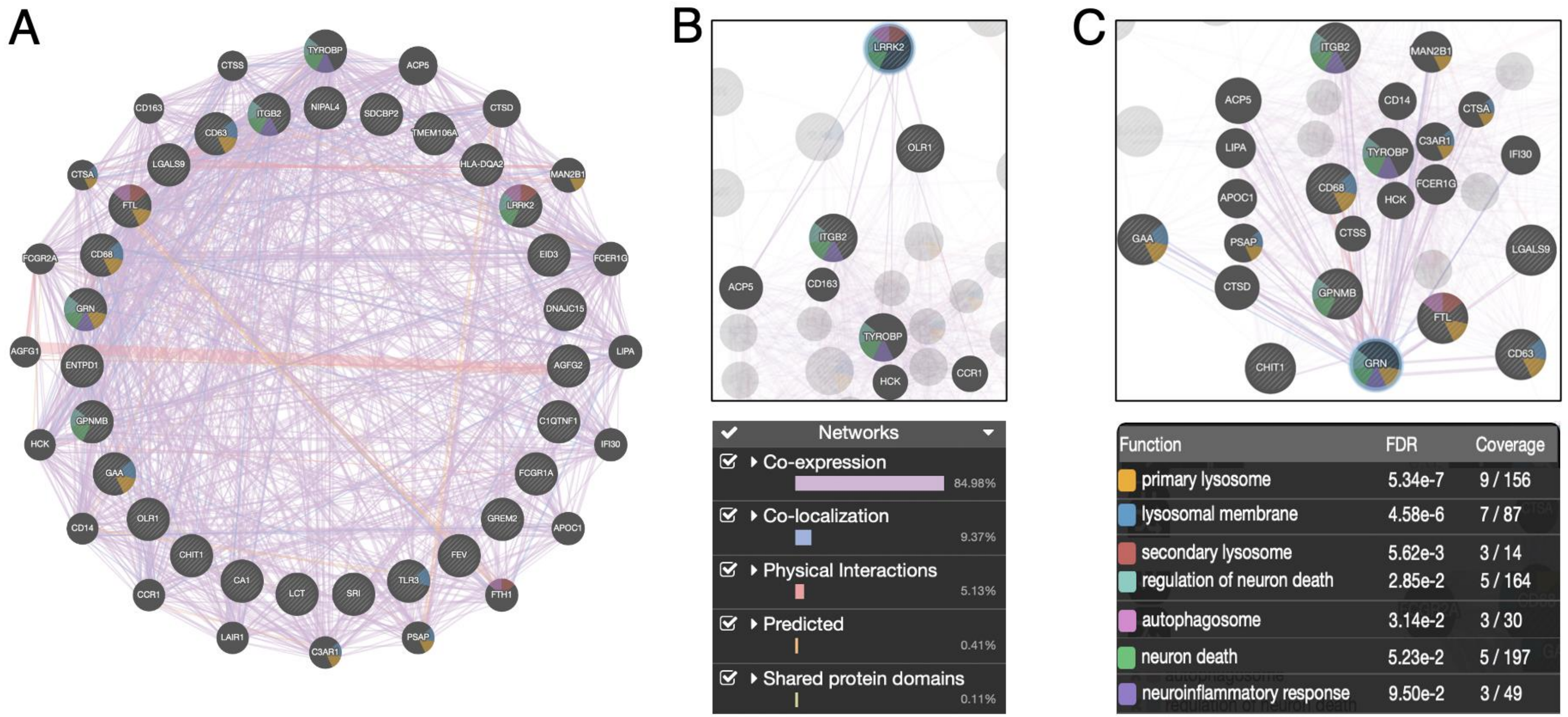


**Supplementary Figure 4: Interaction pathways of LRRK2 associated proteins.** (A) GeneMANIA identified 20 affiliated genes with network interaction and pathways of 26 genes and *LRRK2*. (B) LRRK2 interactions. (C) GRN & GPNMB interactions.
